# Quantifying the relationship between sub-population wastewater samples and community-wide SARS-CoV-2 seroprevalence

**DOI:** 10.1101/2022.04.28.22274086

**Authors:** Ted Smith, Rochelle H Holm, Rachel J Keith, Alok R Amraotkar, Chance R Alvarado, Krzysztof Banecki, Boseung Choi, Ian C Santisteban, Adrienne M Bushau-Sprinkle, Kathleen T Kitterman, Joshua Fuqua, Krystal T Hamorsky, Kenneth E Palmer, J Michael Brick, Grzegorz A Rempala, Aruni Bhatnagar

**Affiliations:** Christina Lee Brown Envirome Institute, School of Medicine, University of Louisville, Louisville, KY 40202, USA; Division of Epidemiology, College of Public Health, The Ohio State University, Columbus OH 43210, USA; Department of Mathematics and Information Science, Warsaw University of Technology, Warsaw Pl. Politechniki 1, Poland; Division of Big Data Science, Korea University, Sejong, South Korea/Biomedical Mathematics Group, Institute for Basic Science, Daejeon, South Korea; Center for Predictive Medicine for Biodefense and Emerging Infectious Diseases, University of Louisville, Louisville, KY 40202, USA; Department of Medicine, School of Medicine, University of Louisville, Louisville, KY 40202, USA; Department of Pharmacology and Toxicology, School of Medicine, University of Louisville, Louisville, KY 40202, USA; Westat, Inc., Rockville, MD 20850, USA; Division of Biostatistics, College of Public Health, The Ohio State University, Columbus, OH 43210, USA

**Author notes:** Correspondence to: Prof Aruni Bhatnagar, Christina Lee Brown Envirome Institute, School of Medicine, University of Louisville, Louisville, KY 40202, USA. Joint first authors (Ted Smith, Rochelle H Holm).

## Abstract

**Background:** Wastewater-based epidemiology is a promising approach but robust epidemiological models to relate wastewater to community prevalence are lacking. Assessments of SARS-CoV-2 infection rates have relied primarily on convenience sampling, which does not provide reliable estimates of community prevalence because of inherent biases.

**Methods:** From August 2020 to February 2021, we conducted a serial stratified randomized samplings to estimate the prevalence of anti-SARS-CoV-2 antibodies in 3,717 participants, and weekly sampling of community wastewater for SARS-CoV-2 concentrations in Jefferson County, KY. With the use of a Susceptible, Infected, Recovered (SIR)-type model, we obtained longitudinal estimates of prevalence and compared these with wastewater concentration, using regression analysis.

**Findings:** Model analysis revealed significant temporal differences in epidemic peaks; the average incidence rate based on serological sampling in some areas was up to 50% higher than health department rates based on convenience sampling. The model-estimated average prevalence rates correlated well with wastewater (correlation=0·63). In regression analysis, a weekly unit increase in wastewater concentration of SARS-CoV-2 corresponded to an average increase of between 1-1·3 cases of SARS-CoV-2 infection per 100K residents.

**Interpretation:** Publicly available health department incidence rates vastly underestimate true community incidence and wastewater has a high potential to provide robust estimates of community spread of infection.

**Research in context:** *Evidence before this study:* Administratively reported clinical case rates of coronavirus disease 2019 (COVID-19) infected individuals are biased due to a wide range of factors from testing access to concerns about missing low and non-symptomatic and self-tested individuals. Wastewater estimates offer an alternative to support community monitoring based on fecal shedding of the virus but are difficult to interpret when compared with the available public health data sets of infection rates. We examined all English literature until February 24, 2022, on Web of Science and PubMed with the terms [“seroprevalence” or “antibody”] AND [“COVID-19” or “SARS-CoV-2”] AND [“wastewater”]. We identified six studies. None of these studies considered randomized COVID-19 community anti-SARS-CoV-2 antibody testing paired with wastewater data.

*Added value of this study:* The study demonstrates how results from serial stratified randomized serological sampling of the community can be used to build a longitudinal model that can interpolate and extrapolate community levels of infection beyond specific testing dates. Such a model correlates well with wastewater concentrations indicating its utility as a surrogate for infection prevalence. The testing data used in the study were collected before wide availability of COVID-19 vaccines and are therefore unique as they are unlikely to include a significant number of false positive results.

*Implications of all the available evidence:* The study demonstrates that convenience sampling obtained data from health department reporting seriously underestimates community-wide prevalence of infection. In contrast, wastewater-based epidemiology may be a faster, cost-effective, and more robust method of estimating the prevalence of viral infections within specific urban areas.

## 1. Introduction

Since early in the pandemic, wastewater sampling has emerged as a rapid, convenient, and economical tool to assess the presence and temporal changes in the concentration of severe acute respiratory syndrome coronavirus 2 (SARS-CoV-2) in communities.^1^ Approximately 34–52% of coronavirus disease 2019 (COVID-19) infected patients shed SARS-CoV-2 in their feces, up to 16 to 27 days from the onset of symptoms,^2^ and can be included in passive, and anonymous, community wastewater monitoring. Thus, wastewater-based epidemiology appears to be a promising new source of community prevalence data but requires a more reliable clinical reference to develop good models. However, due to its limited use prior to COVID-19, there are no established models that relate wastewater data to community prevalence of infection. Although during the COVID-19 pandemic, frequent and wide clinical testing was conducted to estimate the community prevalence of COVID-19, such estimates relied heavily on non-probability or convenience sampling. Although necessary to readily track infection rates in real time, data from such non-probability sampled population are inherently biased and unlikely to provide reliable estimates of the prevalence and incidence of the infection.^3,4^ Moreover, data from testing only individuals with symptoms are unlikely to gauge prevalence, as many infected individuals show no symptoms, and such data are likely to be always enriched in individuals suspecting infections or experiencing symptoms. Therefore, data reported by local health authorities fail to address meaningfully the need for reliable estimates of spatiotemporal infection, and which account for asymptomatic individuals or individual who have not volunteered for diagnostic testing.

Systematic serological surveys with spatiotemporal resolution offer opportunities for infectious disease surveillance.^5^ A systematic assessment of community-wide spread of infection and immunity could be obtained by randomized sampling, and stratified to include individuals of different age, sex, and socioeconomics as well as those living in different geographic areas.^6^ In the COVID-19 pandemic, randomized serological surveys have been conducted at varying scales in the United States across the states of California, Georgia, Indiana, Kentucky, Oregon, and Rhode Island, but with narrow temporal resolution.^4,7–10^ Although such a snapshot measure of cumulative infection in a narrow window of serology is an accurate way to estimate past prevalence of infection in communities, the lack of repeated measurements reduces the utility of such results for relating to wastewater measures. Wastewater results are not cumulative and change over time, and as such discrete wastewater measurements may fail to identify changes in the infection rates or to identify small recalcitrant refugia of infection in discrete geographic hot spots. In contrast, longitudinal stratified sampling could provide robust estimates required to evaluate the fidelity of wastewater measurements.

Previous combined wastewater and serological surveys at a community scale have focused on Hepatitis A and E viruses.^11,12^ Existing SARS-CoV-2 wastewater to community COVID-19 case models have been fit around results obtained from convenience sampling, models which may underrepresent community trends,^13–17^ as well as limited random samples of communities via SARS-CoV-2 in nasal swabs^10^ or blood bank serological surveys.^15,18^ Nonetheless, robust longitudinal estimates of community serology are more appropriate to interpret wastewater measurements.

The purpose of the current study was to evaluate the nature of wastewater measurements of SARS-CoV-2 in relation to community COVID-19 prevalence, adjusted for geographical and demographic variables. To accomplish this, we compared the rates of SARS-CoV-2 infection obtained from serial, stratified random serological samples in conjunction with serial sampling of virus levels in community wastewater. Using statistical prevalence modeling adjusted for both uncertainty in seropositivity measurements as well as for heterogeneity in temporal and spatial epidemic trends, we evaluated the correspondence between wastewater viral levels and estimates of community prevalence obtained through randomized stratified clinical sampling. The simultaneous analysis of these contemporaneous data sets enabled a quantitative comparison of both sampling approaches. We believe that this approach represents the most reliable and economical monitoring and surveillance program for an infectious agent attempted to date.

## 2. Methods

### 2.1 Probability temporal seroprevalence of COVID-19

The study area was the Louisville/Jefferson County metropolitan area in Kentucky (KY), USA, which has a population of approximately 767,000 individuals and represents the largest urban population center in Kentucky. Four probability designed testing waves lasting approximately a week were conducted. The waves were separated by one to three months. Households were sampled using an address-based sampling frame derived from US Postal Service delivery files.^19^ For each wave between 18,000 to 36,000 invitations to participate in the study were mailed to the addresses and the sampled adult was asked to complete a screening interview and schedule an appointment for testing. The response rate was approximately 3%. See Supplement A for details.

### 2.2 Clinical COVID-19 positive case rates

Administrative data pertaining to daily counts of publicly reported COVID-19 infected individuals by street address by date from July 6, 2020, to February 28, 2021, were provided by the Louisville Metro Department of Public Health and Wellness (LMPHW) under a Data Transfer Agreement. We consider the official statistics of the LMPHW as convenience samples. See Supplement B for details.

### 2.3 SARS-CoV-2 (N1 gene) concentration in wastewater

Influent wastewater samples (N=244) were obtained from five water quality treatment plants corresponding to the sampling sectors once to four times per week from August 17, 2020, to February 22, 2021, for the presence and the concentration of SARS-CoV-2 (N1).^20,21^ Some of the sewershed dynamics have been provided in previously published reports.^21–23^ The five subpopulations jointly comprise approximately 97% of the county population. This allowed for the capture and separation of different wastewater regions within the larger county (see Figure 1). See Supplement C for details.

**Figure 1.**
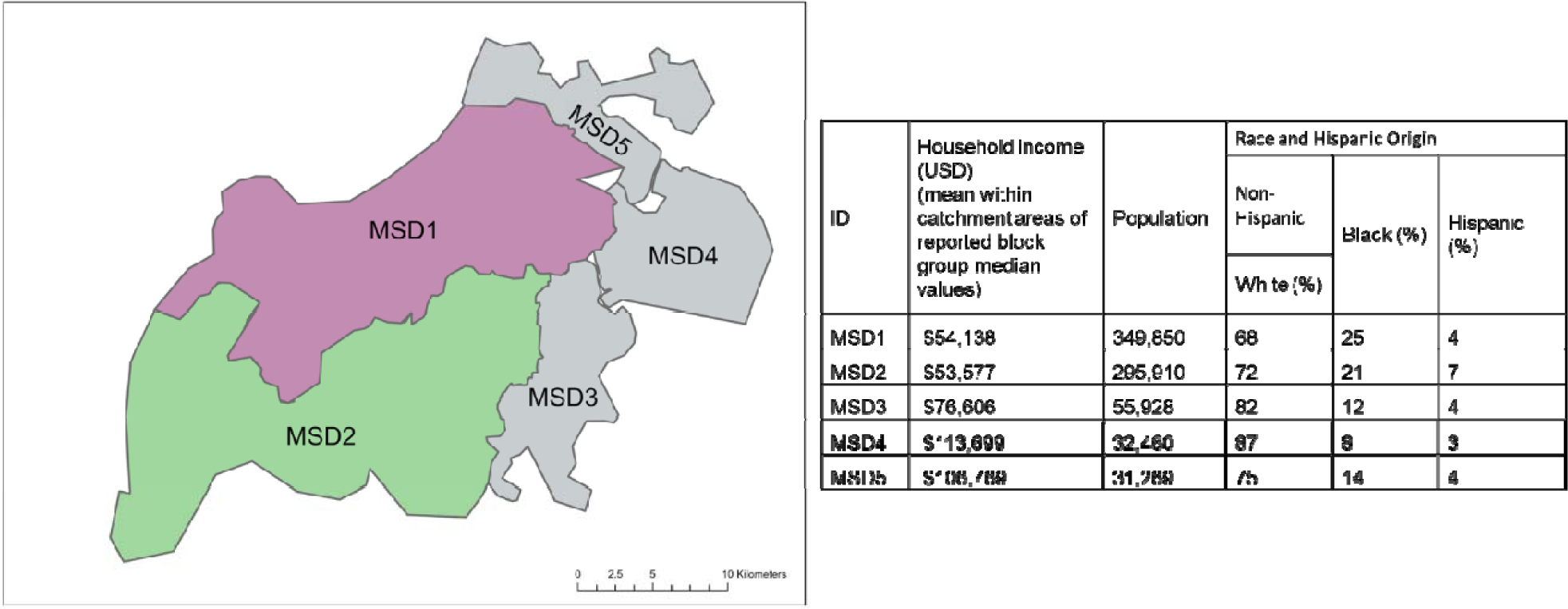
Distribution of wastewater sewersheds in Jefferson County, Kentucky (USA). The adjacent table lists the salient demographic features of each sewershed. Different colors correspond to different spatial strata in the analysis.

### 2.4 Estimating COVID-19 prevalence based on community seroprevalence testing

Serostatus was determined as a qualitative assessment by measuring levels of SARS-CoV-2 spike protein specific immunoglobulin (Ig) G (Spike IgG) antibodies in peripheral blood samples as reported previously by Hamorsky *et al*.^24^ Seroprevalence can detect the antibodies of COVID-19 patients up to 300 days following infection.^25,26^ There was a low number of participants positive for Spike IgG antibodies for the individual areas of MSD3, MSD4 and MSD5 (MSD3-5 N=31 participants positive) (Table 1). To ensure proper balance of the demographic profiles of the subpopulation in the stratified analysis, the data from the three smallest areas were pooled together resulting in three spatial strata.

**Table 1.** Seroprevalence of SARS-CoV-2 by wave and location (95% credibility interval).

|  | Number of participants | Number of participants positive for SARS-CoV-2 Spike protein specific immunoglobulin (Ig) G antibodies | Estimated posterior average seroprevalence per 10<sup>5</sup> people (95% CI) | Estimated posterior average prevalence per 10<sup>5</sup> people (95% CI) |
| --- | --- | --- | --- | --- |
| Overall |  |  |  |  |
| MSD1 | 1998 | 88 | 4702 (2330, 7074) | 78 (1, 155) |
| MSD2 | 1063 | 70 | 6253 (3400, 9105) | 136 (18, 254) |
| MSD3-5 | 621 | 31 | 6231 (3113, 9348) | 174 (14, 334) |
| Other | 35 | 3 |  |  |
| Total | 3717 | 192 | 17186 (12340, 22032) | 388 (175, 601) |
| Wave 1 |  |  |  |  |
| MSD1 | 295 | 10 | 2153 (1651, 2656) | 16 (1, 31) |
| MSD2 | 121 | 3 | 2230 (1691, 2769) | 20 (1, 38) |
| MSD3-5 | 88 | 2 | 2210 (1708, 2712) | 12 (0, 24) |
| Other | 2 | 0 |  |  |
| Total | 506 | 15 | 6593 (5701, 7485) | 48 (21, 75) |
| Wave 2 |  |  |  |  |
| MSD1 | 935 | 22 | 3240 (2109, 4371) | 51 (2, 100) |
| MSD2 | 372 | 15 | 3953 (2151, 5756) | 91 (20, 163) |
| MSD3-5 | 271 | 5 | 3698 (2175, 5221) | 90 (13, 165) |
| Other | 15 | 0 |  |  |
| Total | 1593 | 42 | 10891 (8274, 13508) | 232 (117, 347) |
| Wave 3 |  |  |  |  |
| MSD1 | 480 | 32 | 4461 (2437, 6485) | 106 (1, 211) |
| MSD2 | 342 | 21 | 6003 (3098, 8909) | 192 (38, 346) |
| MSD3-5 | 134 | 3 | 6032 (3120, 8944) | 217 (23, 411) |
| Other | 9 | 1 |  |  |
| Total | 965 | 57 | 16496 (11911, 21081) | 515 (246, 784) |
| Wave 4 |  |  |  |  |
| MSD1 | 288 | 24 | 7342 (2720, 11963) | 119 (0, 238) |
| MSD2 | 228 | 31 | 10416 (5335, 15498) | 222 (18, 425) |
| MSD3-5 | 128 | 21 | 10416 (5335, 15498) | 332 (19, 645) |
| Other | 9 | 2 |  |  |
| Total | 653 | 78 | 28264 (19200, 37328) | 673 (281, 1065) |
| IgG = immunoglobulin G |  |  |  |  |

The model used for estimating prevalence from seropositivity, referred below as SIRT, is a modification of the classical Susceptible, Infected, Recovered (SIR) ecological model of an epidemic^27^ with an additional compartment (denoted T) for seropositive. The tracking of seropositivity status appears necessary as most individuals do not build detectable levels of antibodies until sometime after the infection.^25^ The SIRT model uses a system of ordinary differential equations (ODEs) to describe time evolution of the proportions of susceptible (S), infected (I), removed (R), and seropositive (T) individuals in a large population. See Supplement D for details.

To apply the SIRT model for estimating prevalence, we adapted the ODE-based survival analysis method proposed recently.^28^ Following KhudaBukhsh *et al*., ^28^ we treat ODEs trajectories *S*_*t*,_ *I*_*t*_, *R*_*t*_, *T*_*t*_ as respective probabilities that a randomly selected individual from large population is, at time *t*, susceptible, infected, recovered, or seropositive. In this model, we considered the results of all individual antibody-based tests conducted at time *t* as independent binary variables with probability of a positive test given by *T*_*t*_*\* = T*_*t*_*+(1-spe)(1-T*_*t*_) where *spe* is the specificity level of the diagnostic test (100% sensitivity level of the test is assumed). Given that at time *t, n*_*t*_ individuals are tested with *k*_*t*_ testing positive, the corresponding log-likelihood function is *LL*_*t*_*(θ) ∝ k*_*t*_ *log T*_*t*_*\*+(n*_*t*_ *- k*_*t*_*) log(1-T*_*t*_*\*) where θ* denotes the vector of SIRT model parameters to be estimated. We use the Bayesian Markov chain Monte-Carlo method for estimating *θ*, to properly capture prior information and to account for various sources of uncertainty. With estimated values of the parameters available, we can apply the SIRT model ODEs to calculate average estimated prevalence over time.

### 2.5 Correspondence between estimates of SARS-CoV-2 prevalence from community seroprevalence testing sampling and wastewater measurements

Using contemporaneous wastewater concentration and estimated prevalence, we derived a regression-based correspondence model between the average wastewater concentration and the average prevalence of COVID-19 in Jefferson County both in aggregate and stratified by sewersheds locations. The prevalence rates were calculated based on the SIRT model estimates, with census data and geo-coding techniques used to estimate the actual infection counts by sewersheds. The analysis was based on the linear regression model for prevalence rate and on a negative binomial regression for infection counts.

### 2.6 Ethics

For the seroprevalence and data on COVID-19 infected individuals provided by the LMPHW under a Data Transfer Agreement, the University of Louisville Institutional Review Board approved this as Human Subjects Research (IRB number: 20·0393). For the wastewater, the University of Louisville Institutional Review Board classified this as non-human subjects research (reference #: 717950).

## 3. Results

### 3.1 Seroprevalence and prevalence of COVID-19 in Louisville/Jefferson County

Table 1 shows the number of adults tested and the percent who were positive for SARS-CoV-2 Spike IgG antibodies in the four waves of data collection by the three areas and the SIRT model estimated values of average seroprevalence and prevalence in cases per 100K people. The estimates of average seroprevalence appear to largely follow the pattern of empirical values, increasing over different testing waves, as more individuals first got infected and acquired antibodies to the virus. Note; however, that this is not the pattern of the model-based prevalence estimates presented in the last column. Indeed, the estimates of prevalence indicated by wave four the epidemic was already declining in MSD1 but still expanding in MSD3 to MSD5 that experienced overall higher average prevalence during the testing period than other sewersheds. In Figure 2 we present the model-based prevalence predictions (left panels) as well as the aggregated and stratified model-based seroprevalence fit (right panels). The different prevalence trends both in terms of epidemics peaks sizes and timings in different sewersheds (marked by red lines) are clearly visible in the plots.

**Figure 2.**
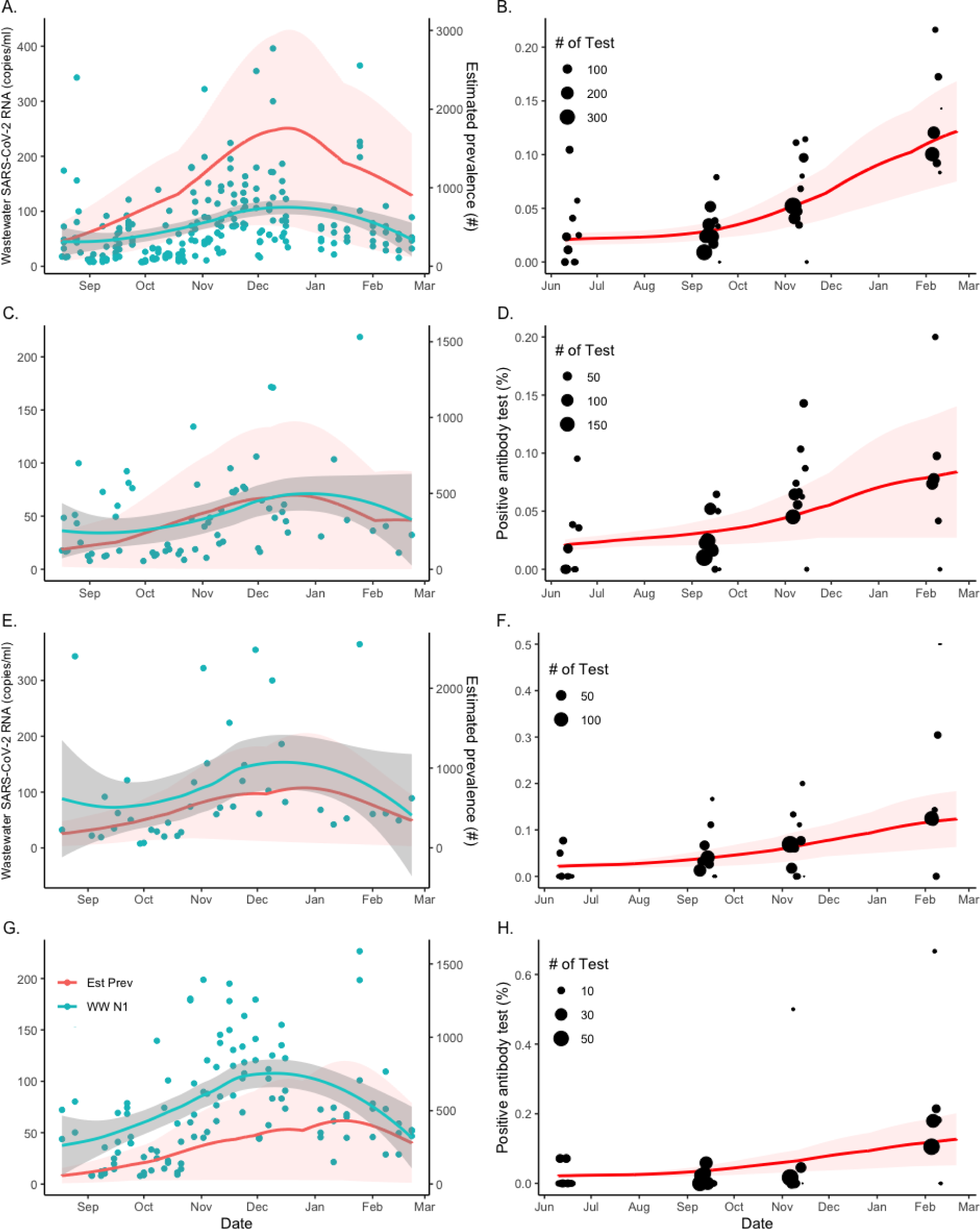
Sampled seroprevalence and wastewater concentrations of SARS-CoV-2 in Jefferson County (Panels A-B) and in different sewersheds (Panels C-H) at indicated dates in 2020-2021. Sewershed areas are stratified as MSD1 (C-D), MSD2 (E-F), MSD3-5 (G-H). **Left (green red) panels**: Temporal changes in the SARS-CoV-2 (N1) concentration in wastewater (discrete blue dots). Solid red line is the model-estimated prevalence. Shaded area corresponds to 95% credibility bounds. **Right (red) panels**: Percent seroprevalence in study participants is shown as black dots indicating sample-size weighted observation, and the red line is the best fit of the model median prediction. Shaded area corresponds to the 95% credible interval.

### 3.2 Correspondence between estimated prevalence and wastewater concentration

SARS-CoV-2 was detected in 90% of the wastewater samples. The stratified serial plots of their mean weekly concentrations are shown in the left panels of Figure 2, superimposed on the corresponding prevalence estimates. The community wastewater concentrations and prevalence estimates show good qualitative agreement over time. To quantify the extent of agreement, we performed two types of Bayesian regression analysis. In the first analysis, we regressed the aggregate and stratified SIRT model estimates of percentage prevalence on the wastewater concentrations via simple linear regression. In the second, we used the Bayesian negative binomial (NB) regression model and regressed the model-predicted prevalence counts on the same set of wastewater abundance. The respective data and model predictions are shown in Figure 3, using weekly aggregated data for better visualization.

**Figure 3.**
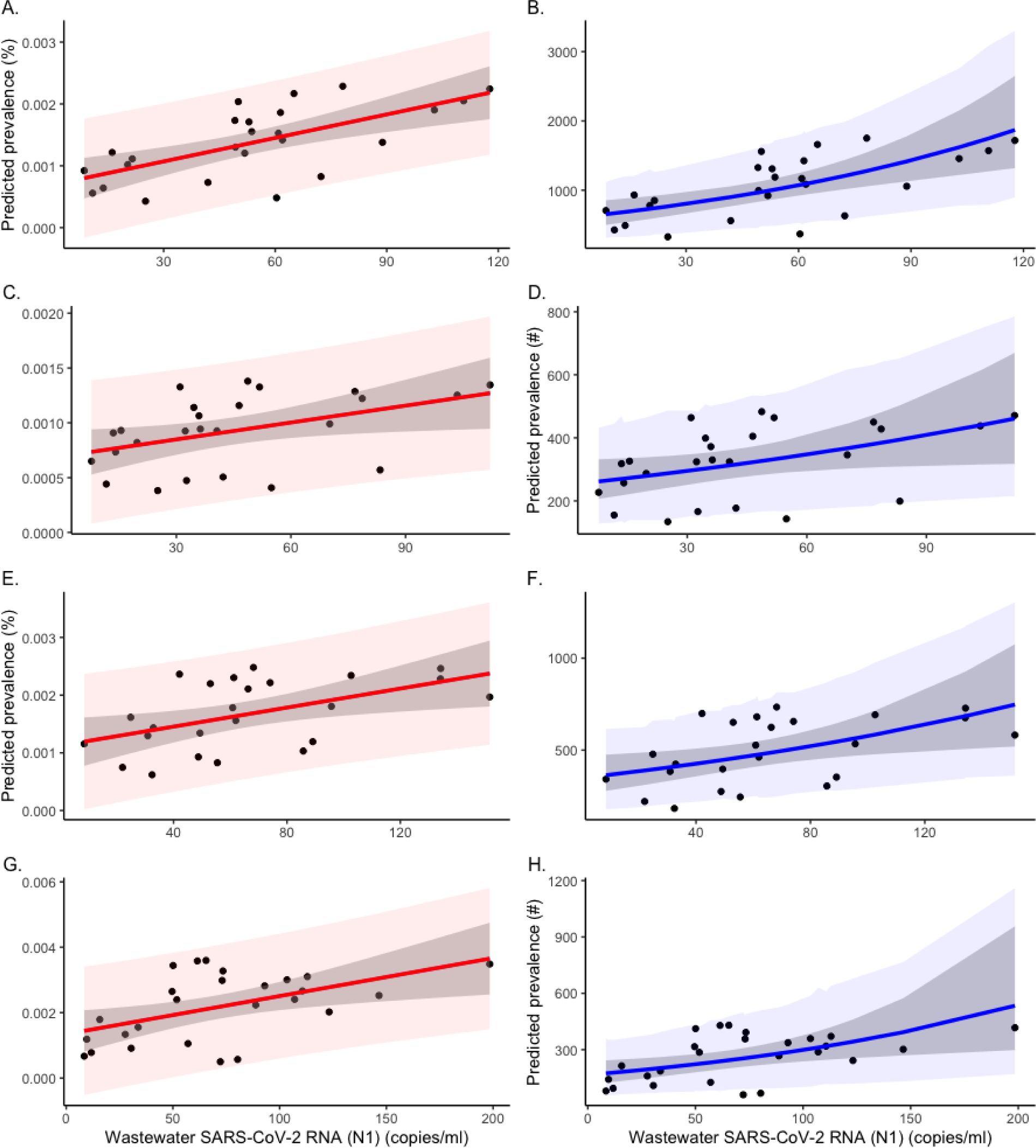
Relationship between predicted weekly prevalence of SARS-CoV-2 infections and wastewater concentration of SARS-CoV-2 in the entire Jefferson County (Panels A-B) and stratified by sewershed areas (Panels C-H). Sewershed areas are stratified as MSD1 (C-D), MSD2 (E-F), MSD3-5 (G-H). **Left (red) panels**: The linear regression of the weekly percent prevalence on weekly SARS-CoV-2 (N1) concentration in wastewater (marked by dots). **Right (blue) panels:** The negative binomial regression of adjusted prevalence (count per 100K people) on weekly SARS-CoV-2 (N1) concentration (marked by dots).

For the pooled data from Jefferson County (top left panel), the results of a simple linear regression model showed a strong correlation of 0·63 (posterior CI = (0·32, 0·84)) between the prevalence of SARS-CoV-2 and average wastewater SARS-CoV-2 concentration. In the aggregate linear regression model (top right panel), the estimated slope coefficient corresponds to relative prevalence increase of 1·27 cases per 100K people (posterior CI = (0·67, 1·88)), for every unit increase in the wastewater concentration. The rates for different watershed areas are listed in Table 2. Similar results were obtained from NB regression, which is more appropriate for directly modeling infection counts. For aggregated data, the NB regression model (top right panel) gives the log-scale regression coefficient to be 0·0097 (posterior CI = (0·00452, 0·151)) corresponding to the prevalence increase of about 1·01 case per 100K people for one unit increase in wastewater concentration. The remaining sewershed zone rates from the NB regression are given in Table 2. All Bayesian regression models are based on flat (non-informative) prior distributions for the model parameters.

**Table 2:**
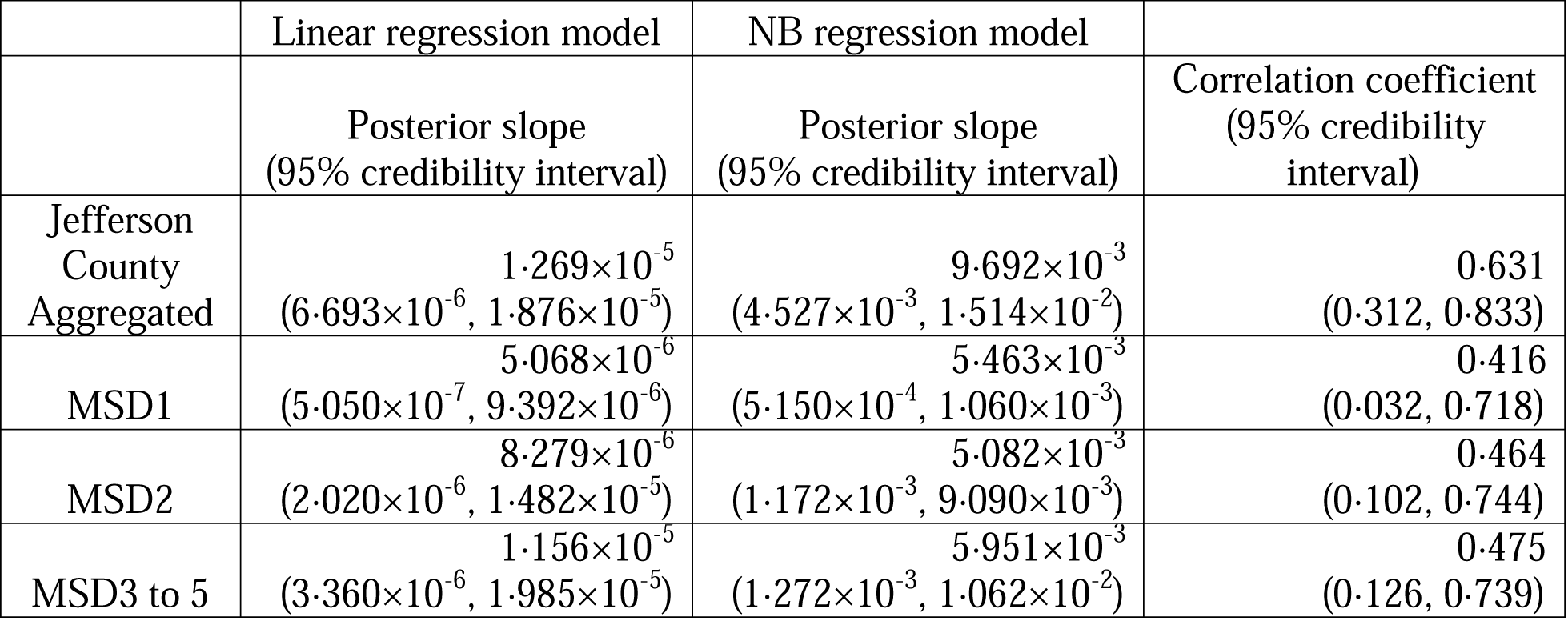
Values of posterior means derived from regression analysis of prevalence versus mean wastewater concentrations (see Figure 3). The posterior means for simple and NB regressions are based on MCMC analysis (see supplemental D material).

### 3.3 Comparison with administratively-reported estimates

In addition to comparing our SIRT-based prevalence estimates with the observed wastewater concentrations of SARS-CoV-2, we have also compared the SIRT-based incidence estimates with publicly reported COVID-19 cases in Jefferson County. The weekly counts were chosen to smooth the administrative reporting variability and weekend reporting delays. The results of our analysis suggest that, as expected, the model-based incidence estimates obtained from the observed seropositivity rates in four waves of testing were significantly higher than the official incidence. The ratio of our model-estimated to officially reported cases was found as, respectively, 1·47, 1·14, 1·48, and 1·80 for the pooled county data and when stratified by sewershed areas MSD1, MSD2, and MSD3 to 5 (Supplement D).

## 4. Discussion

Wastewater-based epidemiology (WBE) has emerged as a rapid, convenient, and economical approach for assessing the prevalence of viral infections in communities. However, WBE estimates lack external validation and it is unclear how well virus levels in wastewater correspond to community-wide prevalence of infection. In this study, we examined the relationship between wastewater SARS-CoV-2 concentration and the prevalence of infection in the community. Rather than using direct official rates, which are subject to bias due to convenience sampling, we estimated the prevalence of infection by repeated measurements of seropositivity in randomized sampling of area population. We developed models for both – estimates of community prevalence and wastewater concentration. The wastewater data correspond well to community prevalence. Combined analyses of these models indicate that one unit prevalence increase in wastewater concentration corresponds to 1 case per 100K people.

A comparison of the administratively-reported COVID-19 cases with our model suggests convenience sampling significantly underestimates the rates of infection in the community. However, the extent of underestimation varied among different sewersheds from a ratio of 1·14 to 1·80. This is similar to the geographical trends in wastewater results for these sewerzones, which are also variable.^22^ Although other research purports a transfer function relating wastewater measures and clinical prevalence is possible,^15,16,18^ it has not been quantified before. Xiao *et al*.^17^ suggests a ratio of wastewater viral copy numbers to reported COVID-19 cases (WC ratio) which changes over time, whereas our model adapts over time but with a consistent transfer function. Moreover, our results do not support a linear relationship between wastewater and prevalence proposed by Layton *et al*.^10^ and Cao *et al*.^13^ Previous convenience seroprevalence studies paired to wastewater may have underreported the correlation, owing to inadequate or no case data from portions of a community, underreporting of at-home COVID-19 antigen rapid self-test results, and reliance on clinical syndromic surveillance. Hoar *et al*.^14^ have suggested that a log10 change in the SARS-CoV-2 viral load in wastewater corresponds to a 0·6 log10 change in the number of new laboratory-confirmed COVID-19 new cases/day in a sewershed. Although their model focuses on predicting new cases and hence is not directly comparable with ours, we find a qualitative agreement between both models.

The utility of SARS-CoV-2 Spike IgG antibodies to estimate community-wide prevalence of infection observed in our study is consistent with previous work.^6^ The use of systematic serological surveys for calibrating wastewater measurements removes much of the selection bias observed in previous relationships.^13–17^ Although seropositivity provides estimates of past infection, an increase in seropositivity over a defined period is likely to be a reliable indicator of the spread of infection. Hence, over the course of the eight-month project, enriched contextual data were provided to city decision makers and stakeholders to inform long-term trends in infection rates as well as the levels of SARS-CoV-2 in local wastewater. The environmental data provided by wastewater sampling when compared with data obtained from in person exams allowed for the development of comprehensive, internally-validated data sets for assessing the trajectory of COVID-19 in the community and for the identification of geographically-defined subpopulations where the virus was lingering. As approximately 97% of the population of Jefferson County, KY uses the sewage system, there measurements of SARS-CoV-2 abundance in the wastewater is largely representative of the community, which, when coupled with a randomized population-based seroprevalence sampling strategy, makes this an ideal dataset for estimating the correspondence between estimates of seroprevalence and WBE. Both our wastewater and randomized seroprevalence have a wide temporal window of capture. While the model was developed from data obtained by using anti-spike protein antibodies in unvaccinated populations, it could be readily modified for vaccinated communities by using anti-N-protein antibodies.

The major strength of this approach is the repeated, randomized sampling design to estimate community-wide changes in seroprevalence paired with frequent wastewater sampling concentration changes, which enabled us to make a mathematical function that models how the inputs relate to each other. To our knowledge, this is the only effort to pair randomized longitudinal seroprevalence with wastewater to-date. The robust coupling of probability-based serological results with wastewater measurements underscores the utility of this approach, not only for cross-validation of each estimate, but also to provide reliable data to support public health decisions. Hence, the findings reported here could form the basis of additional in-depth investigations as it outlines an efficacious approach to follow changes in infection rates in specific geographic areas within a large metropolitan area. Finally, although reported here for SARS-CoV-2 infection, the approach could also be readily extended to the measurement and validation of other viruses and bacteria, as well as other wastewater analytes such as pharmaceutical or xenobiotic metabolites.

## 5. Limitations

Despite its many strengths, this study has some limitations. The shedding rate and duration by COVID-19 infected persons is highly individual.^2^ Both seroprevalence and wastewater are a measure of a portion of the community that has been infected, not necessarily active infections. Our seroprevalence study design only considered an adult population, while wastewater included the entire community population using flush toilets in the county (likely in the range of 2+ years old). Wastewater sampling excluded anyone not connected to the piped infrastructure, such as residents on septic-tanks. We also did not consider the fecal shedding rate in our model. The broader generalization of the specific modeled relationship may be affected by sewer-system dependent factors that are not fully understood.^29^ For example, the wastewater matrix includes complex chemistry and structural specifics which may affect the amount of virus that is recoverable. Replication in other locations would greatly assist in identifying the contribution of such differences.

## 6. Conclusion

In this study, we proposed a novel analytical model to predict disease prevalence of SARS-CoV-2 using systematic retrospective serological surveys with spatiotemporal resolution making a direct comparison of concentrations of viral RNA in wastewater to disease prevalence at a sub-community scale possible. Because we used a community-wide random sampling approach, it is likely that our survey design model has less biased underlying data than previous studies based entirely on convenience sampling, as it was less susceptible to underreporting of community infections and therefore more comparable to the population shedding into wastewater. Our spatial estimates were in good agreement with corresponding data on SARS-CoV-2 wastewater concentrations. Our findings indicate that wastewater data could be used as a surrogate of COVID-19 prevalence as well as the prevalence of other pathogens.

## Data Availability

The seroprevalence and wastewater data used in the study can be accessed at https://github.com/cbskust/Seroprevalene.COVID19. The computer code implementing our model-based analysis will be made available immediately following publication. The code will be shared with researchers who provide a methodologically sound proposal approved by GAR and AB. Proposals should be directed to; requesters will need to sign a data access agreement.

## Contributors

AB, TS, KEP and RK conceived and developed the idea for the study. KEP supervised the serology assays; ABS and KK performed the serology assays, performed quality control assessments, and uploaded the data to the redcap database. TS and RH supervised the wastewater analyses. GAR performed the model-based data analyses, which was supported by CAR, KB, and BC. RHH wrote the first draft of the manuscript with all authors contributing to the interpretation of data and critical revision of the article. MB accessed and verified the data reported in the study. All authors had full access to all the data in the study and had final responsibility for the decision to submit for publication.

## Supplementary material

### Supplement A

#### Probability temporal seroprevalence of COVID-19

Individuals 18 and up were randomly recruited from eight sampling sectors covering the entirety of the county, in total 123,213 adults were invited, and 3,792 participants enrolled. The invited sample included about 16% more minority participants than an equal allocation. Differential response rates are typical in many household surveys, with lower rates of responding associated with high density and non-white households when the primary recruitment method is by the internet. The oversampling of the minority population resulted in a sample of minorities closer to the Louisville/Jefferson County population proportion. The sampling fraction was 3,792/767,419 or 0·49%. At a mobile testing site, nose swabs were acquired to test for COVID-19 infection by PCR and blood was collected for assessing seropositivity. Demographic information including sex, race, age, and address were noted along with vaccination status and location (given as sewerzone).^1^ The team informed each participant of their infection and immunity status by letter. Under the assumption that infection-derived antibodies do not wain to undetectable levels during this analysis the percent positivity of the antibody tests by day acts as a single estimate for seroprevalence on that day.

All households in Louisville/Jefferson County were stratified into 8 sectors roughly proportional to the sector size (population) based on the census block group of the address. In the first four waves of the probability sample (June 2020 to February 2021), a sample of between 2,000 and 3,000 households was selected in each sector (the sampling procedure changed for later waves) using an address list derived from United States Postal Service delivery files.^2^ Each selected household was mailed an invitation to participate in the study in which the sampled adult (18 years or older) was asked to complete an online informed consent, screening, and survey questions, and schedule an appointment for clinical testing. With the mailed invitations, each household of the probability sample population was provided with a unique personal identification registration code to be entered at the time of online registration, thereby allowing the investigators to differentiate between probability and convenience sampling populations. Each household was contacted multiple times to encourage participation. Though a convenience sample was also recruited, the data are not included in the analysis here.

**Figure A1:**
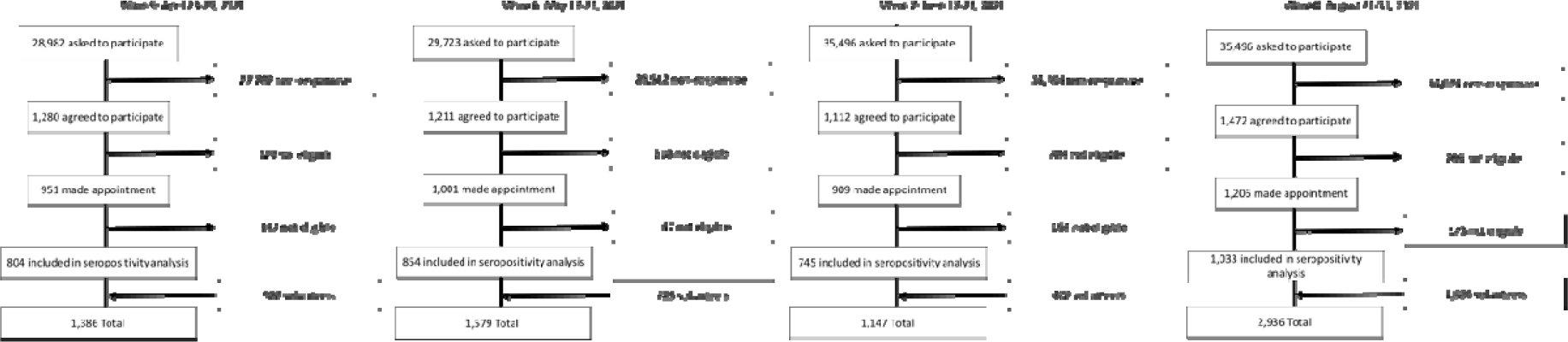
Flow chart of participants in four waves of the Co-Immunity COVID-19 study.

## Supplement B

### Clinical COVID-19 positive case rates

Louisville/Jefferson County reported daily COVID-19 infected individuals by street address by date from July 6, 2020, to February 28, 2021, were provided by the Louisville/Jefferson County Information Consortium and Louisville Metro Department of Public Health and Wellness under a Data Transfer Agreement. Data prior to July 2020 was not reliable. First, locator software was used to automatically assign longitude and latitude coordinates to addresses. For those not able to be automatically assigned, records missing street address information and those with an address outside the county were excluded, and remaining records were manually reviewed. Records with only zip codes for an address were probabilistically assigned to census blocks based on population weighting within each block of that zip code, and records were then assigned the location of the center of the census block. Geocoding was conducted maintaining Health Insurance Portability and Accountability Act (HIPAA), using ArcGIS Pro version 2·8·0 (Redlands, CA). 7-day totals were used to offset effects of clinical testing artifacts such as reporting lags were calculated of new COVID-19 cases based on testing in each of the five-water quality treatment plant zones. In total N= 65,038 publicly reported COVID-19 cases in Louisville/Jefferson County were able to be assigned to the five sewerzones by week.

## Supplement C

### Presence of SARS-CoV-2 (N1 gene) concentration in wastewater

Influent wastewater samples were obtained from five water quality treatment plants corresponding to the sampling sectors once to four times per week for the presence and the concentration of SARS-CoV-2. The five subpopulations jointly comprise approximately 97% of the county population. This allowed capture and separation of the different county wastewater regions, samples (N=244) were collected from August 17, 2020, to February 28, 2021, with the exception sampling was interrupted the last two weeks of December. Samples were collected by the Louisville/Jefferson County Metropolitan Sewer District (MSD) using an automated 24-h time-weighted composite sampler. Samples were transported on ice to the University of Louisville for analysis. SARS-CoV-2 in collected samples was quantified in triplicate by reverse transcription polymerase chain reaction (RT-qPCR) with polyethylene glycol (PEG) precipitation.^1^ Samples were processed within 12 h of collection. Data for SARS-CoV-2 (N1) is reported in copies/ml of wastewater for this work. The threshold value, as recommended by Klymus *et al*.^2^ for N1 assays was 7·5 copies/ml.

**Figure C1:**
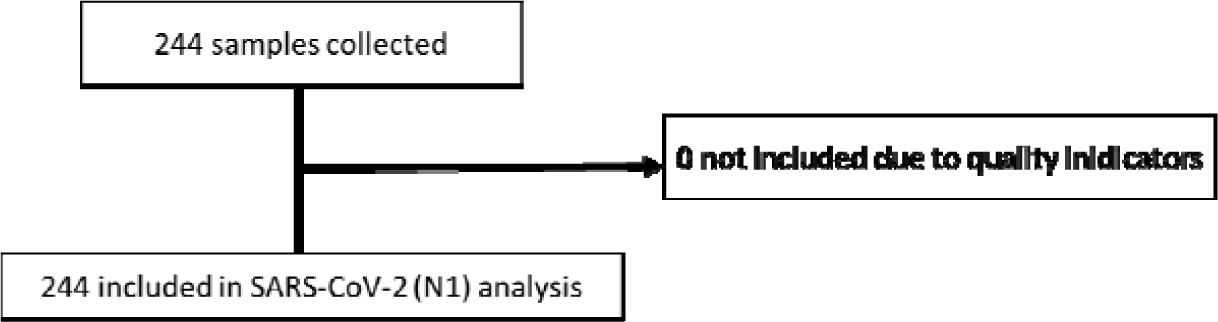
Flow chart of August 2020 to February 2021 of the Co-Immunity COVID-19 wastewater study data.

**Table C1:** Sampling site characteristics in Louisville/Jefferson County Metropolitan Sewer District (MSD)

| Site name | Income<br>(USD\$)<sup>a</sup> | Area<br>(km<sup>2</sup>) | Combined<br>sewer |
| --- | --- | --- | --- |
| MSD1<br>Morris Forman Water Quality Treatment Center<br>(MFWQTC) | 54,138 | 280 | Yes |
| MSD2<br>Derek R. Guthrie Water Quality Treatment Center<br>(DRGWQTC) | 53,577 | 332 | No |
| MSD3<br>Cedar Creek Water Quality Treatment Center (CCWQTC) | 76,606 | 80 | No |
| MSD4<br>Floyds Fork Water Quality Treatment Center (FFWQTC) | 113,699 | 88 | No |
| MSD5<br>Hite Creek Water Quality Treatment Center (HCWQTC) | 106,769 | 67 | No |

## Supplement D

### Systematic assessment of community-wide spread of immunity obtained by randomized seroprevalence and sub-population wastewater sampling: a modelling study

Chance Alvarado, Krzysztof Banecki, Boseung Choi and Grzegorz A. Rempala

#### 1. Details on Data Analysis

The statistical analysis model that allows us to relate serial seropositivity data to both the rates of incidence and prevalence of SARS-CoV-2 in Jefferson County is based on the classical *SIR* ecological model of disease^1^ with an additional compartment, *T*. The rationale behind this compartment is that individuals do not build detectable levels of antibodies for some period after infection.^2^ Below, we refer to this modified model as *SIRT*. Model fitting and parameter estimation for *SIRT* and the related regression models was conducted through the *Stan* statistical modeling platform^3^ integrated into *R* ^4^ via the *Rstan* and *Rstanarm* library.^5^ We specifically used *Stan* to implement a robust hamiltonian Markov chain Monte Carlo (MCMC) algorithm to find posterior probability distributions for model parameters. The final estimates were based on 4000 samples (after burn-in 2000 samples) from converged samplers with Rubin statistic R used to measure convergence based on two independent chains.^5^

##### 1.1. SIRT Model

The equation shown in (1) describes the evolution of the proportions of susceptible (S) infected (I), removed (R), and seroprevalent (T) individuals in a large ecosystem with a small initial fraction of infected. See Brauer^1^ for a brief review. The model equations are

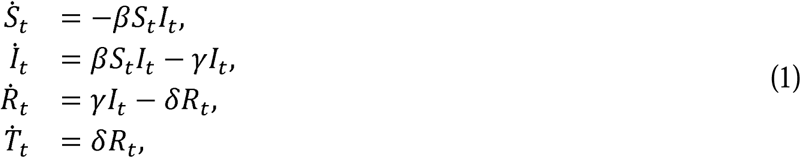

with the initial condition *S*_0_ = 1 *− ρ − ∈ − ψ, I*_0_ = *ρ, R*_0_ = *∈*, and *T*_0_ = *ψ*.

Here, *β* is the rate of infection and *γ* is the rate of recovery. Additionally, *δ* is defined as the rate after recovery at which antibodies build to a detectable level. The *SIRT* model unknown parameters are therefore given by the vector *θ* = (*β, γ, δ, ρ, ∈, ψ*) where all quantities are assumed to be non-negative.

To obtain the serial estimates of incidence and prevalence from the observed seropositivity levels in four waves of testing, we adapt the idea of an ODE-based survival model proposed recently in KhudaBukhsh *et al*.^6^ According to that model, the scaled quantities *S*_*t*_, *I*_*t*_, *R*_*t*_, *T*_*t*_ may be considered as respective probabilities of a randomly selected individual in a large population, being either susceptible, infected, recovered, or seroprevalent at time *t*. Consequently, we consider the results *Z*(*t*) of all individual antibody-based test conducted at times t as independent Bernoulli variables:

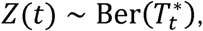

where 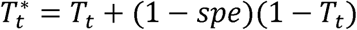 is the specificity adjusted probability of a positive test. For our analysis, we take *spe* = 0.966.

In view of the above, if at time *t, n*_*t*_ individuals are tested with *k*_*t*_ having positive results, the corresponding log-likelihood function is

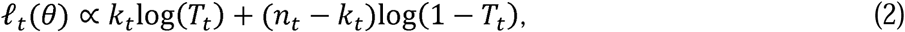

where *θ* = (*β, γ, δ, ρ, ∈, ψ*) is a vector of parameters to be estimated.

Given the testing data at *m ≥* 1 time points *t*_1_, …, *t*_*m*_, we then aim to find parameter values *θ* that maximizes the posterior log-likelihood function

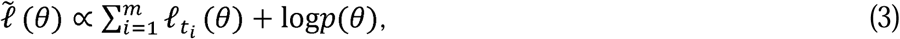

where *p*(*θ*) is the prior distribution on 0 as given in Table 2 in the next section. Hence, we seek the values of *θ* = (*β, γ, δ, ρ, ∈, ψ*) that maximize our posterior log-likelihood function (3). Note that for each parameter combination, the entire system (1) must be solved. Here, this system is solved with a fourth and fifth order Runge-Kutta method.

##### 1.2. Incidence, Prevalence, and Seroprevalence Estimation

Posterior serial estimates of the relative rates of incidence, prevalence, and seropositivity were obtained from the *SIRT* model as the time-dependent vector

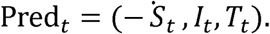

Here (*S*_*t*_, *I*_*t*_, *T*_*t*_) is the family of trajectories of (1) evaluated at the posterior distribution of the vector 0. In practice, the distribution of Pred_*t*_ is approximated by taking a random sample of size m from the converged MCMC sampler. In our case *m* = 1000. For obtaining daily incidence rates (Inc_*d*_) we have used the approximation *S*_*t*_ ≈ *S*_*t+1*_ − *S*_*t*_ and consequently took Inc_*d*_ = *S*_*d*_ − *S*_*d+1*_ where *d* corresponds to a specific day of interest. The estimated prediction counts were obtained by multiplying the rates in Pred_*t*_ by the appropriate population numbers.

##### 1.3. Regression Model for Wastewater Concentration

To relate our SIRT model predictions to serial wastewater measurements of SARS-CoV-2 concentrations, we performed Bayesian linear regression and negative binomial regression based on both the county aggregated data and data stratified by sewershed area. To improve regression model stability, we used cumulative weekly incidence counts from SIRT model as the outcome variable and weekly aggregated average wastewater concentrations as the single explanatory variable. Although the water flow rate adjustment was initially performed, the effect of this adjustment was seen as statistically insignificant, and it was not included in the final model (see Figure S4). In the Bayesian linear regression models, we assigned non-informative priors. Specifically, nob-informative Cauchy distribution was assigned to regression coefficients and non-informative gamma prior was assigned to the dispersion parameter of the error term. In the Bayesian negative binomial regression model, we similarly assigned non-informative normal priors to regression coefficients and non-informative exponential priors to the dispersion parameter. The summary of the posterior estimates of all regression parameters is presented in Table S3 below and the slope parameters summaries are additionally listed in Table 2 in the main body of the paper.

**Figure D1:**
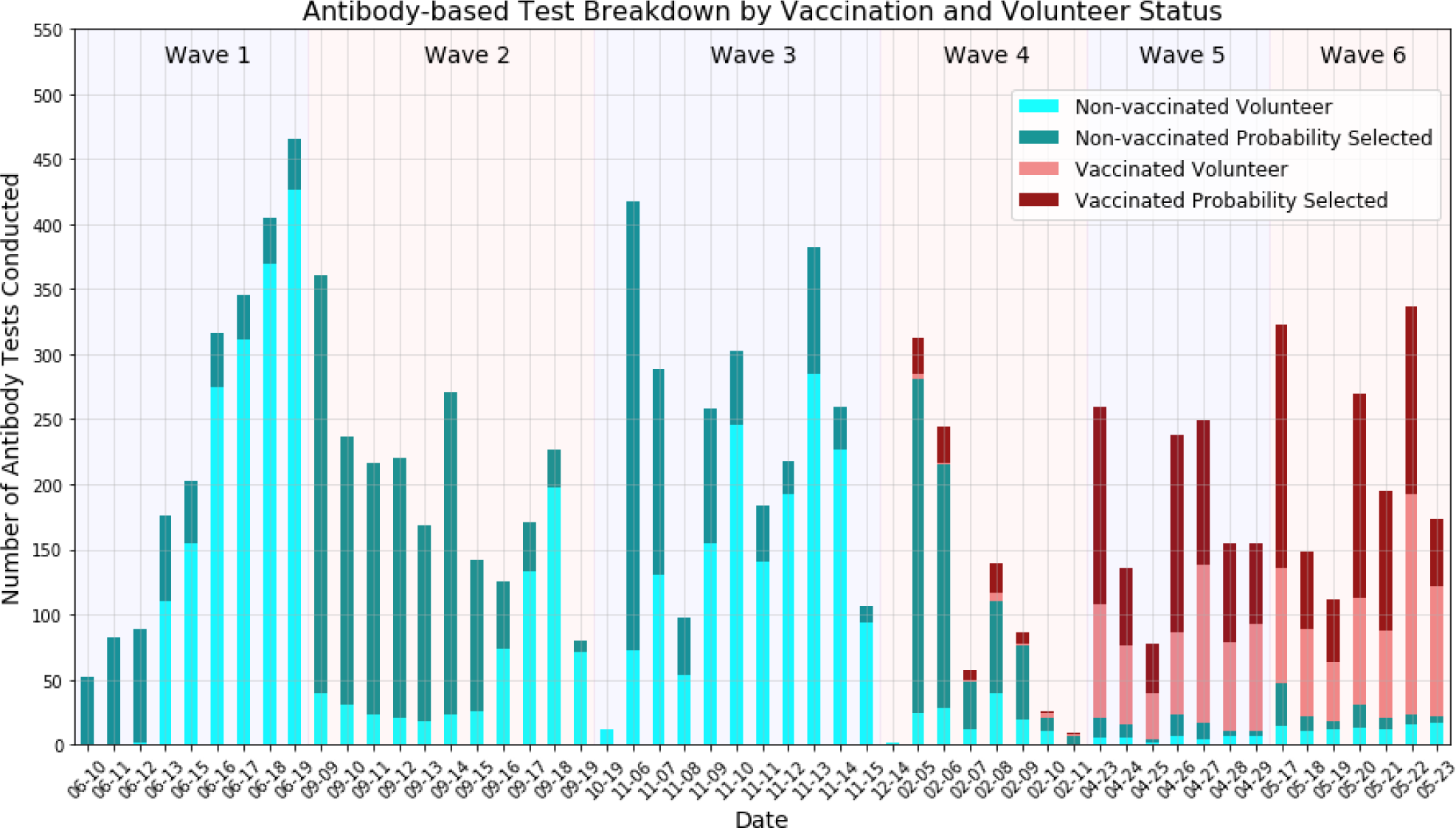
Histogram of antibody-based tests conducted by day - separated by testing waves. Earlier waves (waves 1-4) consist mostly of non-vaccinated individuals while later testing waves (wave 5 and 6) consist of mostly vaccinated individuals. The ratio of volunteer tests to probability selected individuals varies widely between waves.

**Figure D2:**
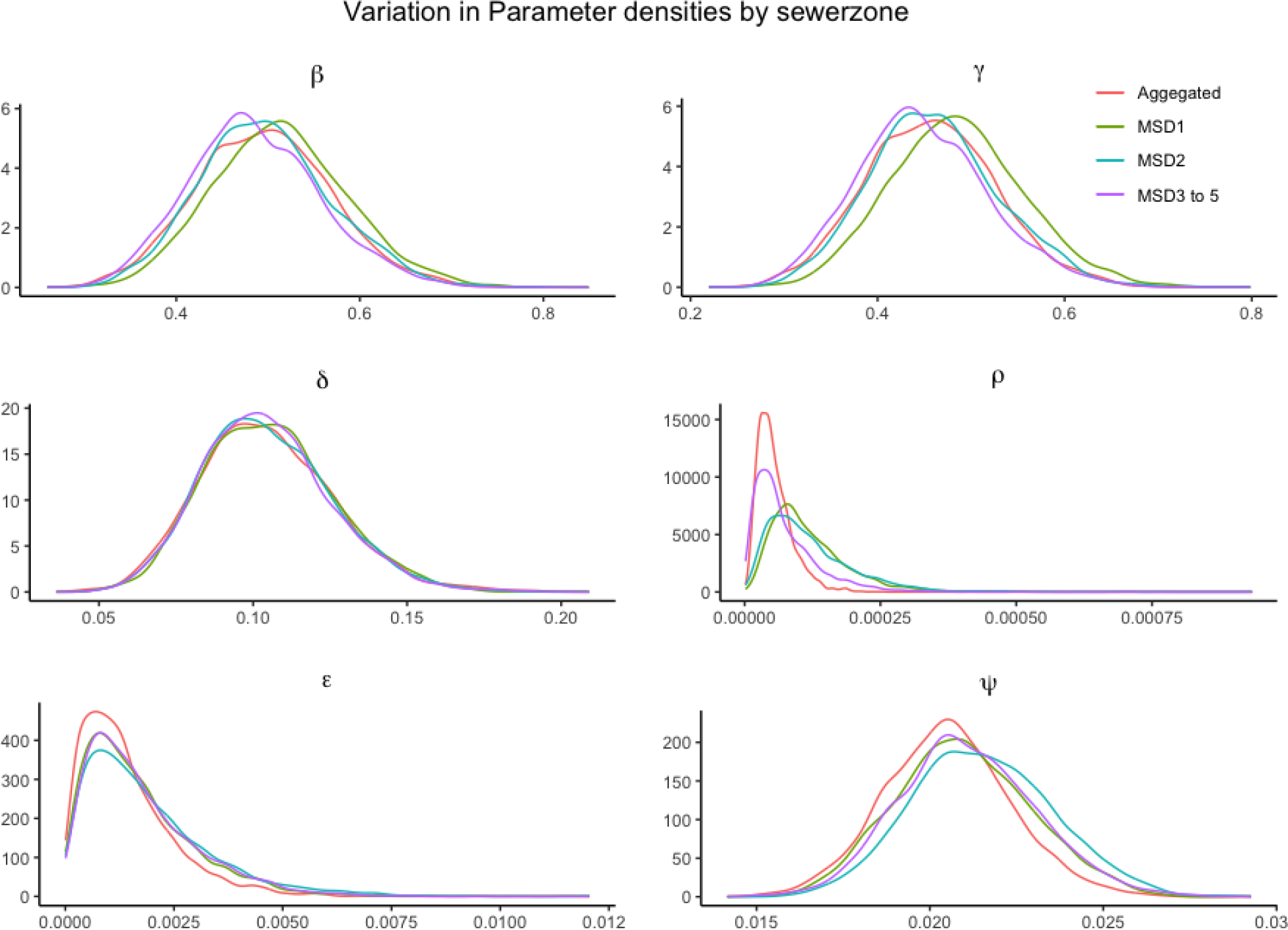
Posterior density of the SIRT model parameters obtained for Jefferson county aggregated and sewershed stratified seropositivity data. The plots are based on Hamiltonian MCMC samples, with 6000 steps and 2000 steps burn in period.

**Figure D3:**
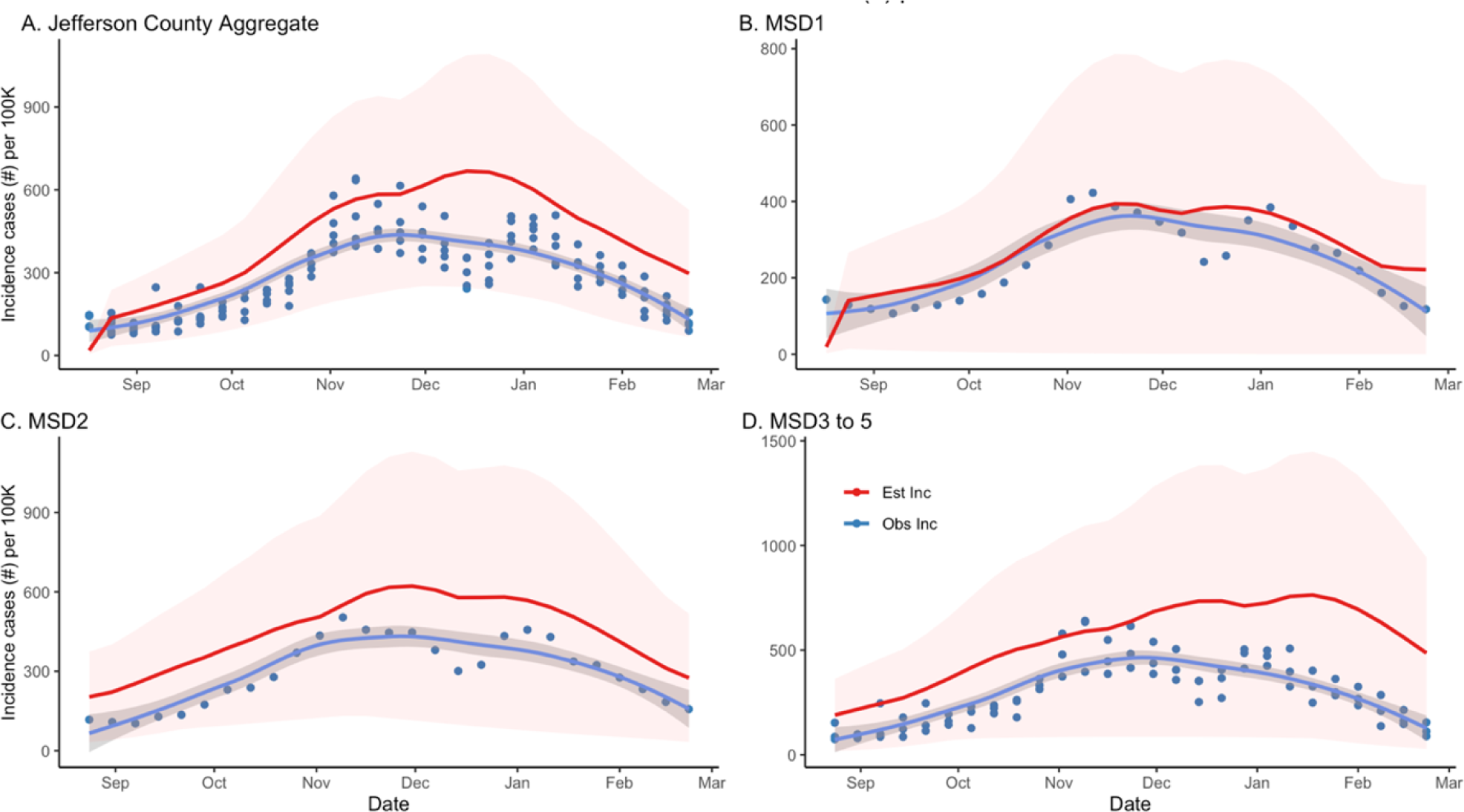
Posterior density and credibility bounds (red curve) of the weekly aggregated incidence rate as predicted by the *SIRT* model compared to the official weekly incidence for Jefferson County (blue dots and trend line) as reported by the Jefferson County Health Department. The panels compare aggregated incidence for Jefferson County (A) as well as incidence stratified by sewershed area (B-D). The model plots are based on Hamiltonian MCMC samples, with 6000 steps and 2000 steps burn-in period.

**Figure D4:**
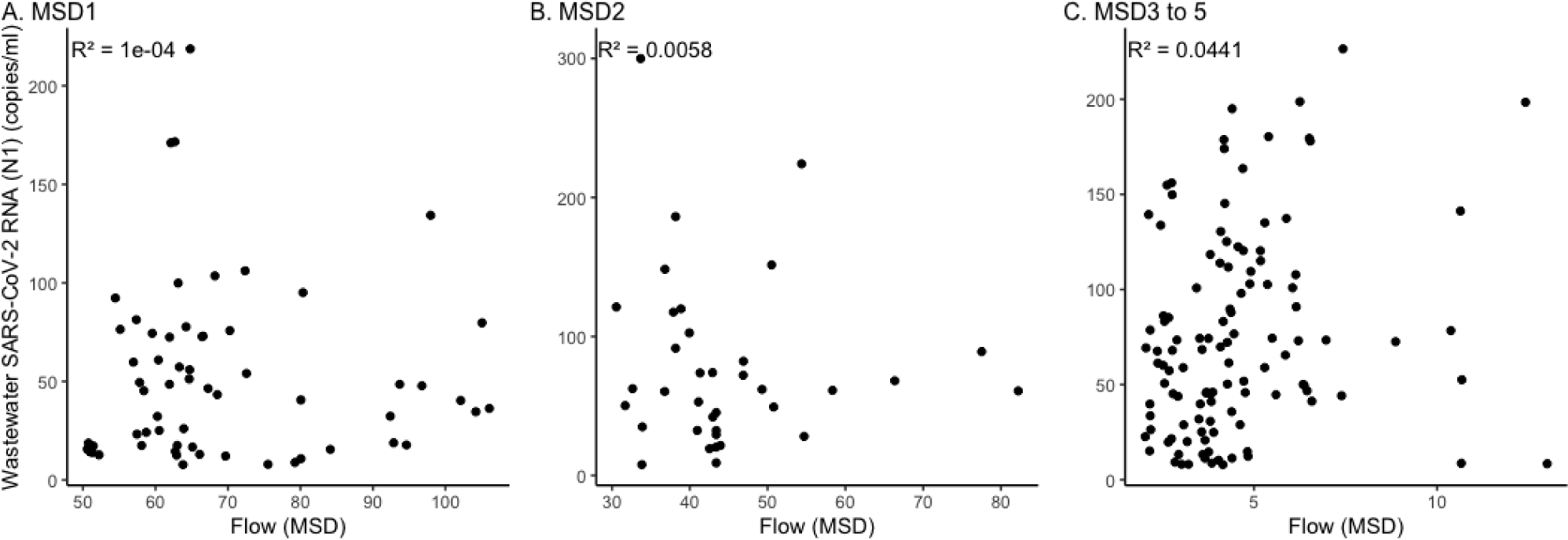
Scatter plots of the sewershed specific flow rates vs average concentrations of wastewater SARS-CoV-2 (N1) (copies/ml). The plots indicate no visible correlation between flow rates and wastewater concentration measurements.

**Table D1:** Posterior mean estimates in the *SIRT* model based on data seropositivity data aggregated across Jefferson County (column 1) and stratified by sewersheds (columns 2-4). The corresponding 95% credibility bounds are provided in parenthesis. The results are based on Hamiltonian MCMC implemented via *rstan* library, with 6000 steps and 2000 steps burn-in.

|  | All Jeff. County | MSD1 | MSD2 | MSD3 to 5 |
| --- | --- | --- | --- | --- |
| $\beta$ | 0.4955<br>(0.4444, 0.6457) | 0.5137<br>(0.4633, 0.6676) | 0.4973<br>(0.4475, 0.6467) | 0.4854<br>(0.4359, 0.6380) |
| $\gamma$ | 0.4571<br>(0.4078, 0.6006) | 0.4849<br>(0.4363, 0.6346) | 0.4614<br>(0.4134, 0.6045) | 0.4480<br>(0.4000, 0.5962) |
| $\delta$ | 0.1045<br>(0.0894, 0.1504) | 0.1054<br>(0.0903, .1497) | 0.1045<br>(0.0895, 0.1496) | 0.1043<br>(0.0897, 0.1481) |
| $\rho$ | 5.47e-05 (3.14e-05,<br>1.35 e-04) | 1.17e-04 (6.77e-05,<br>2.96e-04) | 1.17 e-04<br>(6.01e-05, 3.09e-04) | 7.29e-05<br>(3.16e-05, 2.27e-04) |
| $\epsilon$ | 1.41e-03<br>(6.39e-04, 4.22e-03) | 1.68e-03<br>(7.53e-04, 4.70e-03) | 1.88e-03<br>(8.17e-04, 5.65e-03) | 1.74e-03<br>(7.96e-04, 4.82e-03) |
| $\psi$ | 0.0205<br>(0.0193, 0.0243) | 0.0210<br>(0.0196, 0.0250) | 0.0215<br>(0.0201, 0.0257) | 0.0211<br>(0.0198, 0.0252) |

**Table D2:** The prior distribution specifications for the SIRT model. All parameters were given Gamma prior distributions, with hyper-parameters (*a, b*) with values as listed in the table.

| Gamma (a,<br>b) | $\beta$ | $\gamma$ | $\delta$ | $\rho$ | $\epsilon$ | $\psi$ |
| --- | --- | --- | --- | --- | --- | --- |
| **A** | 40 | 7.85 | 25.41 | 3.1 | 1.79 | 112 |
| **B** | 28 | 61.4 | 239.25 | 11206.96 | 854.29 | 5112.49 |

**Table D3:** The summary of the Bayesian regression result. Both linear regression and negative binomial (NB) models have intercept and slope coefficients. a is the standard deviation of the error term of linear regression and dispersion being a parameter of negative binomial regression.

| Zone | Parameters | Linear regression model | NB regression model |
| --- | --- | --- | --- |
|  |  | Posterior mean (95% credibility interval) | Posterior mean (95% credibility interval) |
| Jefferson County Aggregated | Intercept | 6.937e-04<br>(3.331e-04, 1.052e-03) | 6.3957<br>(6.0746, 6.7088) |
|  | Slope | 1.269e-05<br>(6.693e-06, 1.876e-05) | 9.692e-03<br>(4.527e-03, 1.514e-02) |
| | $\sigma$/dispersion | 4.527e-04<br>(3.419e-04, 6.122e-04) | 7.8021<br>(4.0324, 12.6204) |
| MSD1 | Intercept | 6.967e-04<br>(4.654e-04, 9.278e-04) | 5.5208<br>(5.2533, 5.7843) |
|  | Slope | 5.068e-06<br>(5.050e-07, 9.392e-06) | 5.463e-03<br>(5.150e-04, 1.060e-03) |
| | $\sigma$/dispersion | 3.093e-04<br>(2.330e-04, 4.178e-04) | 8.6179<br>(4.7278, 14.0739) |
| MSD2 | Intercept | 1.121e-03<br>(6.166e-04, 1.595e-03) | 5.8512 (5.5613, 6.1483) |
|  | Slope | 8.279e-06<br>(2.020e-06, 1.482e-05) | 5.082e-03<br>(1.172e-03, 9.090e-03) |
| | $\sigma$/dispersion | 0.521e-04<br>(4.022e-04, 7.485e-04) | 8.9519<br>(4.6814, 14.7946) |
| MSD3 to 5 | Intercept | 1.352e-03<br>(6.602e-04, 2.060e-03) | 5.1087<br>(4.7225, 5.5075) |
|  | Slope | 1.156e-05<br>(3.360e-06, 1.985e-05) | 5.951e-03<br>(1.272e-03, 1.062e-02) |
| | $\sigma$/dispersion | 9.317e-04<br>(7.224e-04, 1.241e-03) | 4.4665<br>(2.4454, 7.1933) |

